# MaveMD: A functional data resource for genomic medicine

**DOI:** 10.1101/2025.11.15.25336228

**Authors:** Abbye E. McEwen, Jeremy Stone, Malvika Tejura, Pankhuri Gupta, Benjamin J. Capodanno, Estelle Y. Da, Sally B. Grindstaff, Nick Moore, David Reinhart, Ashley E. Snyder, Andrew B. Stergachis, Lea M. Starita, Douglas M. Fowler, Alan F. Rubin

## Abstract

Variants of uncertain significance (VUS) undermine genetic medicine implementation because they have an unknown relationship to disease and cannot be used for clinical decision-making. While evidence from multiplexed assays of variant effect (MAVEs) can help resolve VUS, major barriers prevent routine clinical use, including data fragmentation and assay calibration. To address these challenges, we present MaveMD (MAVEs for MeDicine), a new interface for the MaveDB database that displays clinical evidence calibrations, provides intuitive visualizations, integrates with ClinVar and ClinGen, and exports clinical evidence compatible with ACMG/AMP guidelines. MaveMD currently contains 476,076 variant effect measurements curated from 82 MAVE datasets spanning 39 disease-associated genes, enabling classification of 75% of ClinVar VUS and 62% of future variants in these genes. MaveMD is designed to support and facilitate future data generation efforts and the use of MAVE evidence in clinical practice, thereby reducing the VUS burden and improving genetic medicine outcomes.

## Introduction

Clinical interpretation of genetic variants is challenging, often because of variants of uncertain significance (VUS) that cannot be used for diagnosis or to guide clinical decision-making. The widely-used American College of Medical Genetics and Genomics (ACMG) and Association for Molecular Pathology (AMP) variant classification framework integrates evidence including population frequency, familial segregation, case-control studies, computational predictions, and functional assays to classify variants as pathogenic, likely pathogenic, VUS, likely benign, or benign^1^. Each piece of evidence can support pathogenicity or benignity and is weighted as very strong, strong, moderate, supporting, or stand-alone. The final variant classification is determined by combining evidence using a point-based system^1,2^. If the point total does not meet the threshold for likely pathogenic or likely benign, the variant is classified as VUS. Up to 40% of individuals undergoing genetic testing receive VUS results^3–5^, disproportionately affecting individuals of non-European ancestry^6,7^.

Data from multiplexed assays of variant effect (MAVEs) are a powerful source of evidence for classifying genetic variants and resolving VUS^8^. These high-throughput experiments simultaneously characterize thousands of variants in a single assay. Each variant receives a functional score from the assay, generating a comprehensive variant effect map for the target gene^9–12^. Incorporating MAVE data into variant classification workflows allows reclassification of ∼55% of VUS across well-studied genes^8,13^, and can play a major role in reducing classification disparities for individuals from non-European ancestries^14^. Although MAVE-derived evidence is valuable for clinical variant classification, its use has been constrained because of barriers to discovering and accessing MAVE data, as well as the lack of necessary expertise amongst clinicians to evaluate data quality, assay utility, and evidence strength^15–17^.

MAVE data has been difficult to access because it is fragmented across supplemental tables of publications, websites maintained by individual laboratories, gene-specific databases, general repositories like GitHub and Zenodo, and the MaveDB community database. This dispersion creates an untenable burden for clinical laboratories compounded by data loss as supplemental tables, websites and databases become unavailable over time^18–22^. Even if clinical users can locate MAVE data, they need key metadata to evaluate its clinical utility and inform its use, including information about the model system and experiment design, the molecular phenotype assessed, and the types of variants the assay detects (e.g., splicing, dominant negative, loss of function, or gain of function). Additionally, clinicians need to calibrate the data, a process that transforms MAVE-derived functional scores into evidence for clinical variant classification. Calibration relies on clinical control variants with previously-established pathogenic or benign classifications from sources like ClinVar^23^ to serve as benchmarks for determining evidence strength^24^. Many MAVE datasets have been calibrated, but these evidence assignments are often also buried in supplementary tables and therefore are not easily discoverable or usable. Moreover, multiple calibration methods are available, including methods that assign variant-specific evidence strength^25,26^ and the ClinGen-specified OddsPath method^24^, a useful quantitative metric that reflects an assay’s ability to distinguish between benign and pathogenic variants.

To promote data discoverability, MaveDB has been established as the community database for MAVE data with 2,752 total datasets, including variant effect measurements in 713 human genes, 487 of which have a known relationship to human disease. In addition to variant functional scores and basic information about the assay target, MaveDB stores text metadata designed to meet the needs of data scientists and researchers, including a short description of the assay, an abstract, and abbreviated methods^27,28^. However, MaveDB lacked features needed by clinical users, including individual variant search, standardized human genomic coordinate mapping, clinically-oriented metadata, and clinical evidence strength for individual variants. Thus, even if clinical users downloaded a dataset from MaveDB, they would then need to merge information from diverse resources before using the MAVE data (**Figure 1A**).

**Figure 1:**
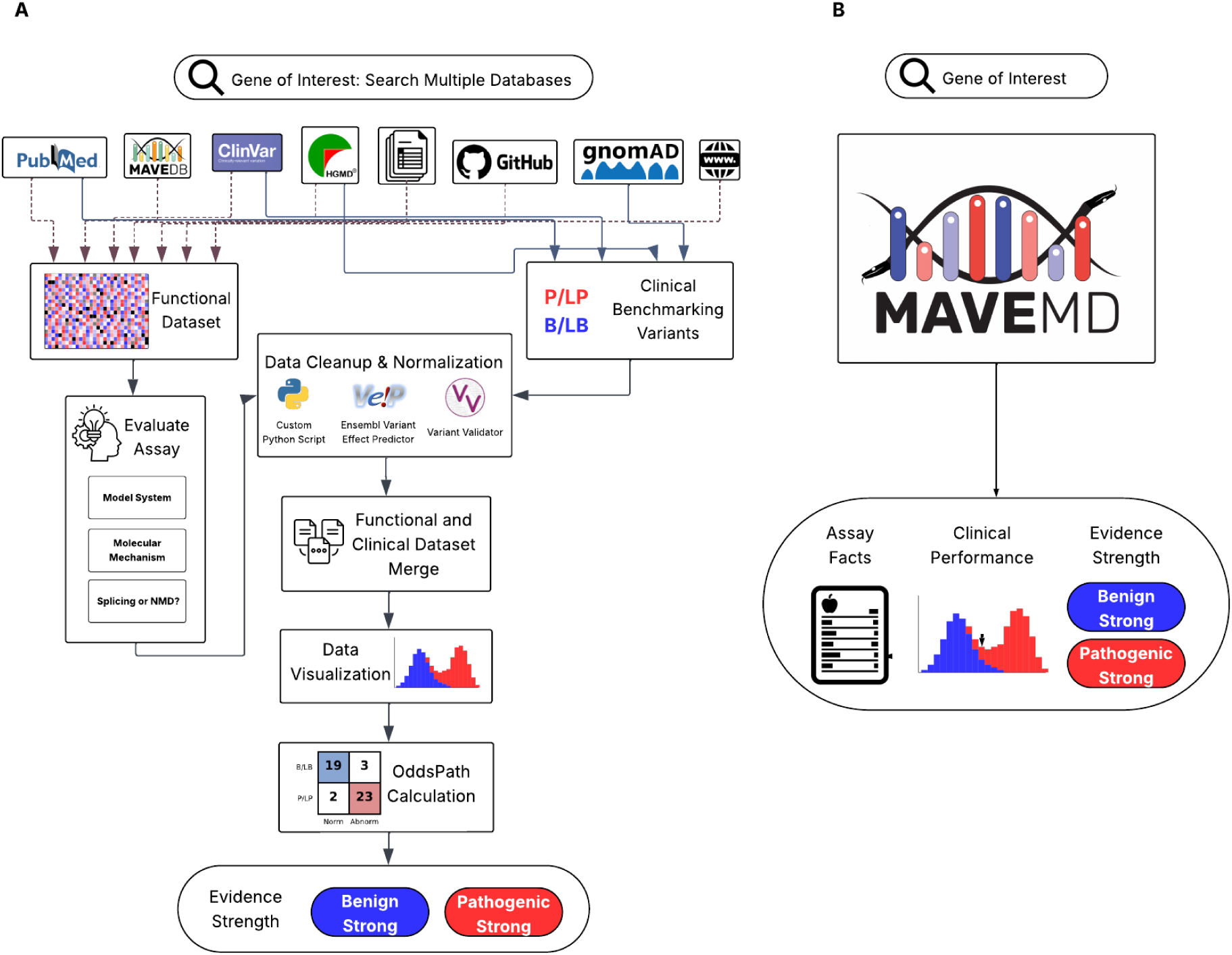
Comparison of workflows for using MAVE data in a clinical context. A) Currently clinical users must retrieve data from multiple databases, perform data cleanup and normalization, merge MAVE datasets with clinical control variants, assess clinical performance, and calibrate the functional/MAVE data to determine evidence strength. (B) The streamlined MaveMD interface displays the results of those steps in a single platform, providing direct access to key assay metadata, visualization of clinical performance, and evidence strength classifications in an accessible, clinician-friendly format.

The clinical utility of MAVE data depends on understanding the relationship between the specific assay molecular phenotype (*i.e.,* what the assay is measuring) and the gene-disease relationship being evaluated. An assay must measure a molecular function directly relevant to the disease pathophysiology to provide meaningful evidence. Therefore, clinicians must determine the molecular phenotype measured, the variant types the assay reliably detects (*e.g.*, loss-of-function, gain-of-function, and dominant negative), and if an assay can detect splicing variants or variants predicted to undergo nonsense mediated decay (NMD). Without this key information, even well-calibrated MAVE data may lead to incorrect variant classifications if applied to inappropriate gene-disease contexts, underscoring the importance of displaying easily understandable assay metadata. Furthermore, MAVE datasets have grown in scale, with some assessing more than 10,000 variant effects^29^, and newer calibration methods are sufficiently complex^26^ that a centralized repository is needed to provide calibrated data to clinical laboratories.

To bridge this gap and bring functional assay data into medicine at scale, we present MaveMD (MAVEs for MeDicine), a resource built on MaveDB and informed by surveys of clinical users to help clinicians discover, evaluate and use MAVE data (**Figure 1B**). MaveMD includes a new variant search function powered by ClinGen Allele IDs to ensure accuracy and portability between information systems^30^. We also developed and implemented a standardized, clinically-relevant metadata model based on existing minimum information standards^31^, and adopted modern Global Alliance for Genomics and Health (GA4GH) standards to power sophisticated API-based data integration^32,33^. MaveMD links MAVE functional scores to known pathogenic and benign clinical control variants from ClinVar, and exposes evidence strength assignments from two calibration methods^24,26^. The clinical interface is accessible via REST API for programmatic access or the web front-end with standardized and intuitive visualizations allowing users to evaluate assay results, explore key metadata, and examine calibrations. We populated MaveMD with an initial set of 82 curated and calibrated MAVE datasets with 476,076 distinct variant effect measurements for 39 clinically-relevant genes, including 12 actionable genes on the ACMG secondary findings list^34^. This includes 251,573 (53%) variant effect measurements with evidence strength assignments from the ClinGen-recommended OddsPath method^24^ and 452,304 (95%) from the research-use-only ExCALIBR method^26^, and 475,199 (99.8%) having evidence strength from either or both methods. The curations and calibrations currently in MaveMD enable reclassification of 75% of ClinVar VUS and 62% of future variants in these genes, demonstrating a scalable approach to overcoming the VUS challenge^35^. Lastly, MaveMD is integrated with the Impact of Genetic Variation on Function (IGVF) Consortium’s Catalog, providing access to MAVE data as it is generated^36^. Thus, MaveMD builds upon MaveDB by adding extensive metadata curation, evidence calibration, and visualizations designed to support routine integration of functional evidence into variant classification workflows.

## Methods

### Data curation and selection criteria

To establish a comprehensive collection of clinically relevant MAVE datasets, we performed systematic curation of published functional studies. We identified candidate datasets by searching MaveDB, querying ClinVar for publications citing foundational MAVE methodology or clinical assay calibration papers^13,24,37^, and soliciting expert recommendations from the genetics community. To ensure focus on genes with sufficient evidence for clinical interpretation, we only included genes with moderate or greater disease associations as determined by either the ClinGen Gene-Disease Validity Working Group classifications or the GenCC database^38^.

A multidisciplinary curation team led by a molecular genetic pathologist (A.E.M.) and including a senior graduate student (M.T.) and genetic counselor (P.G.) met weekly to review curations, resolve ambiguities, and ensure consistency. Systematic metadata was extracted across six key domains encompassing over 180 individual fields. Dataset identification and availability metadata included gene identifiers (HGNC symbols and IDs), publication details (PMID, year, first author), data locations (MaveDB URNs, supplemental tables, repositories), and accessibility status. Assay design parameters captured experimental methodology (arrayed vs. pooled, saturation vs. targeted variants), model system specifications (organism, cell line or yeast details), library strategies (variant types, mutagenesis approach, delivery method), phenotypes measured, and molecular processes investigated. Technical performance metadata included replicate structure, variance analysis between replicates, and comparisons with other functional assays. Functional scoring included methods used for calculating variant effect scores and associated statistics. Score classification described methods for assigning functional classifications and assay thresholds for each functional class. Clinical performance metrics captured the use and source of pathogenic and benign clinical control variants, evidence strength assignments per ACMG/AMP guidelines (OddsPath), and performance statistics (sensitivity, specificity, PPV, ROC-AUC). This comprehensive metadata collection ensured standardized data capture across all curated datasets and directly informed the design requirements for MaveMD.

### Data wrangling and standardization

The heterogeneous nature of published MAVE data necessitated extensive standardization before datasets could be submitted to MaveDB, which underlies MaveMD. Source data, including variant functional scores, were collected in diverse supplemental table formats (Excel, CSV, TSV, PDF) with inconsistent column naming conventions, variant nomenclature, and score representations. We developed a systematic pipeline to transform this information into MaveDB-compatible formats.

For each curated dataset, we performed manual inspection to identify the data’s structure, including the variant representation format (amino acid substitutions, nucleotide changes, mixed nomenclature, custom notation), score columns and their meanings (raw scores, normalized scores, confidence intervals, standard errors), replicate structure when present, and any additional metadata columns requiring preservation. Converting variant descriptions to the HGVS-based format^39^ used by MaveDB was the most complex part of the process. Source datasets employed various nomenclature systems: single-letter amino acid codes, three-letter amino acid codes, nucleotide positions without reference sequences, genomic coordinates in multiple genome builds, and laboratory-specific custom notations. Following standardization and deposition into MaveDB, all datasets were mapped to the human reference genome using sequence alignment as previously described^40^.

### Data counting and summarization

Datasets in MaveDB and elsewhere are counted as unique combinations of a functional assay and assay target. In MaveDB, this is tabulated at the level of the experiment record. For cases where the same raw sequencing data was analyzed in multiple ways to produce different sets of variant effect measurements, these were not considered unique datasets.

After data wrangling and standardization, the 82 curated datasets encompassed 476,076 variant effect measurements deposited in MaveDB. This count reflects all assay-level measurements, where unique genetic variants tested in multiple assays contribute multiple counts to the total. After collapsing variants measured in multiple assays, the dataset contained 268,862 unique variants (**Supplemental Table 1**). In this manuscript, counts reported at the unique variant level are based on this collapsed dataset, while counts at the variant effect measurement level are taken from the complete set of assay-level measurements.

### Interface prototype development and assessment

To inform the development of the MaveMD interface, we utilized findings from a comprehensive needs assessment survey^16^. The survey captured responses from 190 genetics professionals between February and June 2024, who evaluated a prototype MAVE data dashboard featuring ∼4,000 *BRCA1* variants. Participants evaluated eight specific interface features using Likert-scale questions, assessing each component’s potential utility for clinical variant interpretation. The dashboard included a histogram displaying MAVE functional score distributions colored by ClinVar significance with functional thresholds, OddsPath calculations, ACMG/AMP evidence codes, variant identifiers, functional consequences, scores, and associated errors.

### Software implementation

MaveMD is built on the MaveDB open source codebase and shares many key components^28^. Briefly, the software is implemented in Python and JavaScript using FastAPI and Vue.js, respectively, and a Postgres database. MaveDB and MaveMD source code is available under the AGPL-3.0 license (see **Data and code availability**). They are deployed on Amazon Web Services as a series of Docker containers and other services (**Supplemental Figure S1**).

## Results

### Understanding MaveMD requirements

Since MaveDB’s launch, clinical users have consistently requested features such as variant search that would help them leverage MAVE data in their practice. We surveyed 190 respondents, predominantly laboratory medical geneticists (23%) and variant review scientists (23%), using a mockup MaveMD interface with different proposed features: metadata about how the assay was performed, functional classification thresholds (e.g., loss-of-function or functionally normal), standardized variant identifiers, a visualization comparing pathogenic and benign control variants to assay results, and calibrated evidence strength using ACMG/AMP v3 evidence codes^1,16^.

Response to the mockup was overwhelmingly positive, with strong support for all the proposed features^16^. However, while 94% of respondents preferred having access to standardized ACMG evidence codes (PS3/BS3), only 68% stated they would directly use these codes if displayed. This gap revealed that clinical users prefer transparency and want to review both the evidence codes and the underlying supporting data before making decisions. These findings translated into clear requirements for MaveMD: robust mapping of MAVE variants to the reference genome, variant search functionality, inclusion of metadata essential for clinical interpretation, and presentation of calibrations with underlying clinical control variants.

### Contextualizing MAVE data for clinical application

Inspired by the results of our survey, we systematically curated 82 clinically relevant MAVE datasets across 39 disease-associated genes^13,25,29,35,41–81^. Our curation data model included all metadata outlined in the MAVE minimum information standards^31^ augmented with additional fields critical for clinical use, such as the molecular mechanism measured by the assay, variant consequences detected by the assay, and whether the assay could detect splicing variants or variants that would be subject to NMD in a physiologic context. Ultimately, each dataset underwent comprehensive metadata curation across six domains: dataset identification, assay design parameters, technical performance metrics, functional classifications, score calculations, and clinical performance statistics (**Supplemental Table 2**).

To understand the coverage of our MAVE data, we analyzed the overlap between variants characterized in our curated datasets and existing clinical resources. Of the 268,862 unique total variants across all curated MAVEs, 11.55% (31,065) were present in ClinVar, with 4.25% (11,417) having confident clinical classifications (pathogenic/likely pathogenic or benign/likely benign; **Supplemental Table 1**).

Curated datasets reflected diverse experimental approaches, contributing to the difficulties clinicians face when trying to evaluate assays. Arguably the most essential challenge is understanding the type of assay being performed, and this diversity emphasizes the importance of organizing and displaying interpretable metadata (**Figure 2A**). Of the 82 datasets we curated, six combined multiple dissimilar functional assays and were excluded. We found that assays were evenly split between those that measured overall protein function and those that measured a specific function. Assay types included cell fitness (54%), reporters (41%), and direct protein function (5%), with assay types further split into subtypes to help guide application of each dataset.

**Figure 2:**
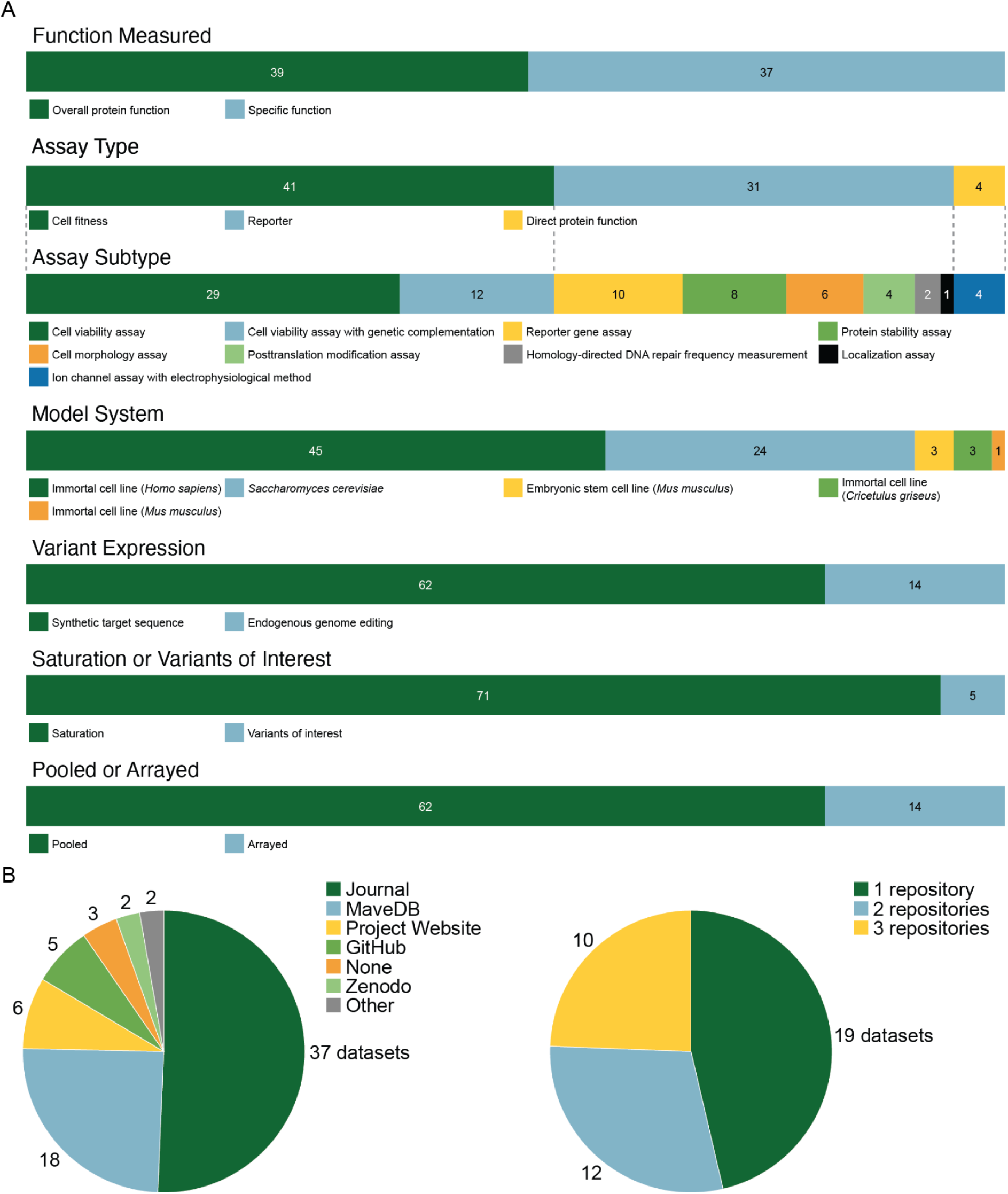
Summary of curated clinically-relevant MAVE datasets. A) The stacked bar plots show the proportion of datasets split across several metadata categories. The legend for each bar is shown below. B) The pie chart demonstrates the location of MAVE dataset deposition at the time of publication.

The assay systems also differed substantially across our curated datasets, with most experiments performed in various mammalian cell lines (68%) and the remainder in yeast cells. Variants were mostly generated using cDNA-based approaches that required introduction of a synthetic target sequence (82%), with the remaining minority using endogenous genome editing techniques such as saturation genome editing. The overwhelming majority (93%) of datasets used saturation mutagenesis, measuring the effects of all possible variants across a region or gene. Of the assays we curated, most were pooled assays, as expected for deep mutational scanning and other MAVE methods (82%), but a few used arrayed formats for technical or historic reasons.

As part of curation, we ensured that each dataset was uploaded to MaveDB. Of the 42 publications analyzed, 17 (41%) already had data deposited, 17 (41%) only had data in supplemental tables on journal websites, and one (2.4%) could only be found on a project-specific academic website, highlighting the discoverability challenges facing clinical users (**Figure 2B**). One curated dataset was associated with links that were no longer functional but available through other sources, and one dataset could not be curated because the only data source was a non-functional website. Likewise, two datasets were excluded because they were in PDF files that could not be easily converted.

For MAVE data to be translated into functional evidence, clinical control variants must be compared to functional classifications derived from variant effect measurements. While most datasets provided a functional classification for each variant or included information such as clearly defined score intervals (55%), the remaining studies did not. Additionally, only a minority of datasets included clinical evidence strength calculations (26%) with 25 using the OddsPath method and four using a variant-specific log-likelihood ratio method.

### MaveMD clinical interface implementation

In developing the MaveMD interface, we were guided by three key priorities: variant search functionality, representation of metadata essential for clinical interpretation, and presentation of calibrations with underlying clinical control variants^16^. Variant search presented significant challenges due to the diversity of transcripts, variant identifiers, variant formats, and coordinate systems. To address this complexity, we utilized the ClinGen Allele Registry, which provides and maintains universal identifiers for genetic variants^30^. We first implemented an automatic mapping process that determines the corresponding human genomic variant for every variant in MaveDB that comes from a human sequence, regardless of the assay system ^40^, allowing us to create or retrieve associated ClinGen Allele IDs from the Registry, as well as display annotations from other resources such as gnomAD (**Figure 3**).

**Figure 3:**
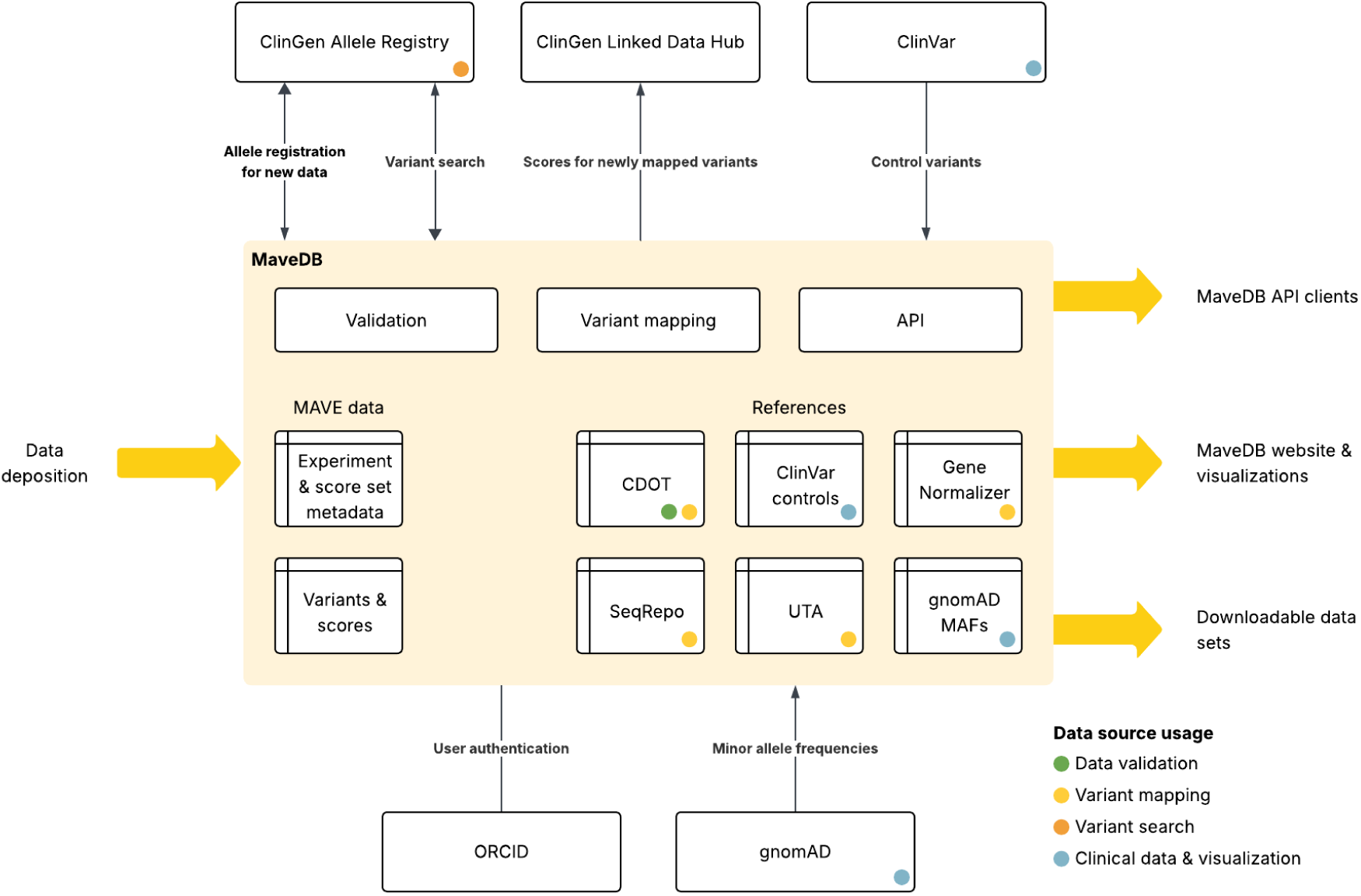
Data flow for clinically-relevant datasets in MaveDB. This schematic shows the various data sources and services used by MaveDB and MaveMD as part of the mapping and annotation process. Sources of reference information include the cdot transcript resolver (https://github.com/SACGF/cdot), classified variants from ClinVar, Gene Normalizer for resolving ambiguous sequences (https://github.com/cancervariants/gene-normalization), SeqRepo for storing a local collection of sequences^82^, the Universal Transcript Archive (UTA) for storing aligned transcripts (https://github.com/biocommons/uta/), and population minor allele frequencies from gnomAD^83^.

Having mapped variants and obtained ClinGen Allele IDs, we were able to implement two types of search designed to support commonly-used variant nomenclatures from clinical reporting systems. Users can search for variants with standard HGVS strings or through a structured “fuzzy search” interface that requires a gene symbol, variant type (protein or cDNA), position, and reference and alternate alleles (**Supplemental Figure S2**). Search results display available MAVE datasets and provide direct access to a purpose-built clinical dashboard, including the option to choose between different functional datasets when multiple measurements are available (**Supplemental Figure S3**). The top panel displays variant identifiers and external links, including ClinGen Allele ID and ClinVar Variant ID, enabling integration and exploration in other clinical resources. Users are also able to link directly to a MaveMD variant from the corresponding variant page in the ClinGen Allele Registry via the ClinGen Linked Data Hub. The variant’s functional consequence is also prominently displayed with a color-coded functional classification.

Upon searching for a specific variant, MaveMD provides a clinical view designed to help users access and interpret MAVE data and clinical metadata for variant classification. We developed a standardized presentation format that transforms MAVE data into actionable information, reducing the workload for clinical users while maintaining scientific rigor and transparency about assay capabilities and limitations. The interface contains three major features: assay fact labels, interactive visualizations displaying functional scores, and evidence calibration information.

The first major feature, assay fact labels, addresses the need for rapid assessment of a functional assay’s relevance to a given clinical application (**Figure 4A**). They display key characteristics including assay type (cell fitness, reporter, direct protein function) and molecular phenotype assessed, model system (*e.g.*, human cells or yeast), variant functional consequences detected (loss of function, gain of function, dominant negative), the ability to detect splicing or NMD variants, and the total number of variants tested. A clinical performance section in each assay fact label provides at-a-glance evidence strength assessments using color-coded ACMG/AMP-style evidence codes and OddsPath values for both normal and abnormal function or a note if OddsPath values are not available. Thus, our assay fact labels highlight critical information for clinicians, allowing rapid comparison between different MAVE datasets using a consistent format, and reveal assays meeting specific clinical needs.

**Figure 4:**
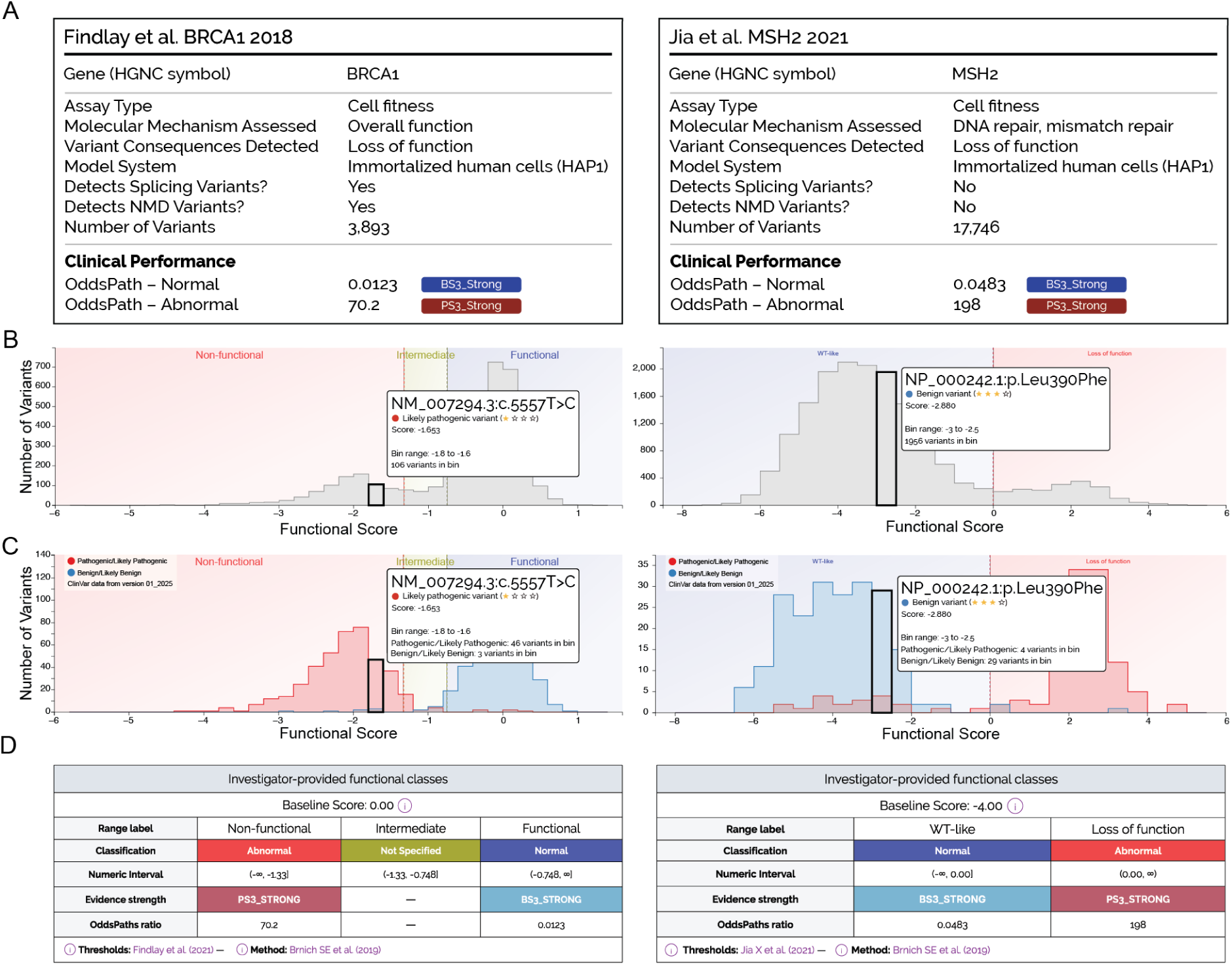
MaveMD data display elements. The MaveMD interface features multiple different visualizations and visual summaries. A) Each dataset has its own assay facts box that summarizes the key details that will help a user evaluate its clinical utility. Where needed, additional references are denoted with a superscript number that links to the full reference on the associated MaveDB or MaveMD page. B) Interactive histograms show the full distribution of variant scores in an assay and the relative position of a selected variant. Vertical bars also denote any evidence- or function-based cutoffs. C) Another interactive histogram displays the distribution of variant scores for variants from ClinVar. This view is intended to help users evaluate the clinical relevance of an assay based on the separation between independently-classified pathogenic and benign variants. Users can choose different snapshots of the ClinVar database. D) OddsPath and the associated evidence codes and score ranges are shown in a table below the histogram.

**Figure 4A** presents two examples: the left panel illustrates *BRCA1* saturation genome editing results that measures overall BRCA1 function related to cell survival and detects both splicing and NMD variants^46^. The right panel shows a cell survival based assay of DNA mismatch repair for MSH2, which, due to its cDNA-based approach, cannot detect variants affecting splicing or causing NMD^56^. This distinction is clinically relevant since a laboratory investigating a putative splice variant needs to quickly identify that cDNA-based experimental methods might yield falsely normal results.

The second major feature facilitates the assessment of assay performance via an interactive histogram displaying the distribution of functional scores across all tested variants. This visualization highlights user-selected variants within the broader context of all variant effect measurements in the assay. In the “Overall Distribution” view, the histogram features clear demarcation of author-specified functional classes with colored backgrounds (for example, blue for “wild type-like” or “normal activity”, red for “loss of function” or “abnormal activity”, yellow for “intermediate”) (**Figure 4B**). This color coding is especially helpful for a dataset like MSH2, as in contrast to most MAVE readouts, lower functional scores actually indicate normal protein activity.

The third major feature provides transparent access to evidence calibrations. By toggling to a “Clinical Controls View”, the interactive histogram displays only the functional scores for classified ClinVar variants and shows OddsPath calculations and evidence strength assignments (**Figure 4C and D**). Variants are color-coded by their ClinVar status, with red for pathogenic/likely pathogenic and blue for benign/likely benign, alongside statistics showing the number of clinically classified variants in each functional score bin. This visualization immediately reveals whether functional scores are concordant with previously-classified clinical variants and highlights potential discordances requiring further investigation. Recognizing that multiple calibration algorithms already exist and continue to be developed, MaveMD allows users to toggle between the results of different methods. For example, we currently provide calibrations from the OddsPath and ExCALIBR methods (**Supplemental Figure S4**)^24,26^.

Users can also download the MAVE datasets, calibrations, and metadata they need to make their own assessments. To preserve the rich contextual metadata required for interpretation, we use the GA4GH Variant Representation Specification (VRS) for describing genomic variants^33^ and the GA4GH Variant Annotation Specification (VA-Spec) for describing assay scores, functional annotations, and clinical evidence strength (https://github.com/ga4gh/va-spec).

## Discussion

Our systematic curation and centralization of MAVE datasets and the new MaveMD interface directly confronts the data dispersion crisis plaguing clinical genetics. The fragmentation we documented, with datasets scattered across supplemental tables, institutional websites, and various repositories, creates a substantial burden for clinical laboratories already managing heavy caseloads. Prior to this work, clinical laboratories faced the daunting task of independently evaluating each functional assay, requiring both deep domain expertise and computational skills often unavailable in clinical settings. By establishing MaveMD as a central hub with persistent identifiers and standardized formats, we eliminate the need for exhaustive multi-platform searches while enabling long-term data discovery and preservation.

Fundamental challenges remain that both limit the current implementation and point toward future solutions. The inconsistent representation of measurement uncertainty across datasets complicates the incorporation of variant-level confidence into clinical interpretation. Much of this heterogeneity is driven by the diverse landscape of scoring methods available to researchers^84^, as well as the distinct sources of error present in different experimental designs. Assembling a harmonized dataset as we have done here is the first step in tackling this systematically by further improving metadata annotations and identifying candidates for large-scale reanalysis.

The most complex unresolved issue is dealing with conflicting MAVE data. When multiple assays for the same variant yield discordant results, clinical users lack a clear framework for reconciliation. Because assays may measure different molecular properties, a system like ClinVar’s transparent display of conflicting interpretations is not suitable. A variant could show normal protein abundance in one MAVE and demonstrate impaired enzymatic activity in another; both results may be correct, but their integration requires sophisticated understanding of disease mechanisms and the relative importance of different molecular functions.

Our current approach of displaying all available MAVE data with associated metadata promotes transparency, but leaves the task of reconciliation to clinical users, and the required expertise may exceed what is reasonable to expect from clinical laboratories. Datasets assembled in MaveMD can be used to train machine learning models generating composite functional scores that leverage the complementary strengths of different experimental approaches, as has been demonstrated for TP53^13,85^. MaveDB already supports multiple score calculations for a single assay, as well as model-based combinations of multiple assays in its data model. While large-scale reanalysis and model training is out of scope for MaveDB’s current role as a data sharing platform, disseminating such results through MaveMD would be straightforward.

While MaveMD significantly improves access to MAVEs for clinical use, important limitations remain in connecting assays to specific gene-disease relationships. Currently, we provide comprehensive assay metadata including molecular phenotypes measured and variant types detected, but clinical users must determine whether a given assay is appropriate. Furthermore, our current implementation displays all pathogenic and benign clinical controls from ClinVar regardless of disease association, as disease annotations are often incomplete, inconsistent, or unreliable. This approach may include variants related to diseases with different molecular mechanisms than the condition of interest, potentially affecting calibration accuracy. Future development should prioritize integration with a centralized, well-curated resource that systematically maps gene-disease relationships to their underlying molecular mechanisms. Such a resource would enable MaveMD to automatically flag which assays are most relevant for specific clinical indications. Until this information is standardized and computationally accessible, clinical users must exercise careful judgment in selecting and applying MAVE evidence, considering both the molecular basis of the disease in question and the specific functional readout of each assay. Our curation experience illustrates the intensive effort required to extract, standardize, and calibrate each dataset, and scaling this manual process to accommodate the accelerating pace of MAVE publications is a fundamental challenge. By reporting crucial insights into what information is essential for clinical interpretation, our expanded data model will help shift this work from post-publication curation to data deposition, while also streamlining the process for data submitters by moving away from free text towards more intuitive structured metadata.

MaveMD and our associated expert curations demonstrate that the primary barriers to implementing functional evidence in the clinic are logistical rather than scientific. By addressing data centralization, metadata and format standardization, and calibration complexity, we have created a foundation for routine use of MAVE and other functional data that we hope will satisfy clinical needs now and into the future. However, challenges around measurement uncertainty, conflicting results, and sustainable maintenance require continued community effort. The long-term success of functional evidence in clinical practice depends on establishing sustainable infrastructure for data generation, data sharing, and platform maintenance. As functional assays become increasingly sophisticated and widespread, parallel advances in computational infrastructure, experimental and analytical methods, and community standards will be essential to realize their full potential for resolving variants of uncertain significance and reducing disparities in genetic medicine.

## Data Availability

All data produced in the present study are available upon reasonable request to the authors or available on https://mavedb.org

Source code is available from https://github.com/VariantEffect/mavedb-api and https://github.com/VariantEffect/mavedb-ui

## Acknowledgments

This work was supported by an NIH NHGRI Advancing Medical Genomics Research award (R01HG013025), and L.M.S. and D.M.F. were supported by NHGRI IGVF grant (UM1HG011969). A.B.S. holds a Career Award for Medical Scientists from the Burroughs Wellcome Fund and is a Pew Biomedical Scholar. A.E.M. was supported by an Early Career Award from the Alex’s Lemonade Stand for Childhood Cancer and RUNX1 Foundation (21-25037) and a Brotman Baty Institute Catalytic Collaborations Grant (CC28). P.G. was supported by the Career Ladder Education Program for Genetic Counselors grant from the Warren Alpert Foundation (WAF-CLEP-PD 10089501-01). This project received grant funding from the Australian Government.

## Data and code availability

MaveDB, MaveMD, and associated online documentation are available at https://mavedb.org. Source code for MaveDB and MaveMD is available at https://github.com/VariantEffect/mavedb-api (for the back-end application) and https://github.com/VariantEffect/mavedb-ui (for the website). The present paper describes version 2026.1.0.

---

**Supplemental Figure S1:**
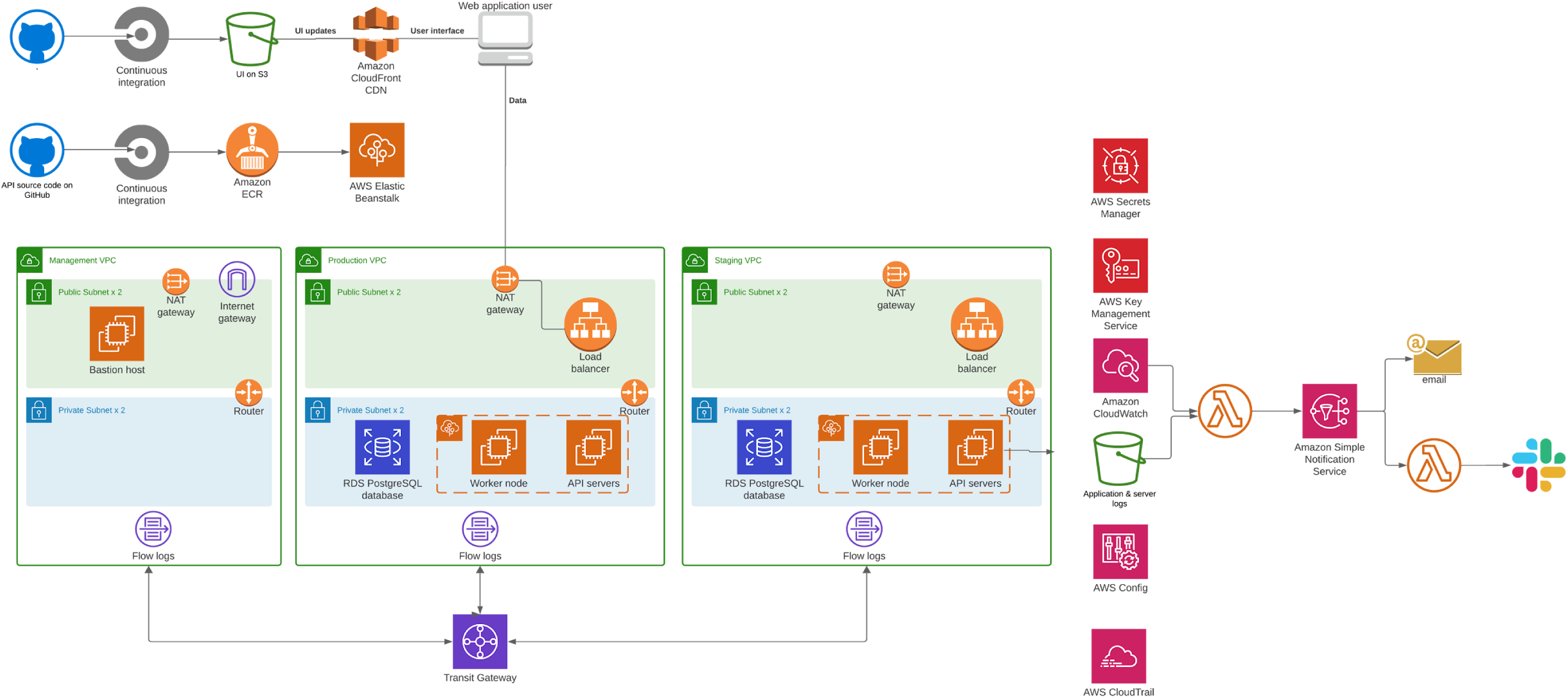
Amazon Web Services (AWS) architecture diagram. MaveDB and MaveMD are hosted in a cloud computing environment provided by AWS. This schematic shows the various processes and containers involved in running, updating, and monitoring MaveDB and MaveMD.

**Supplemental Figure S2:**
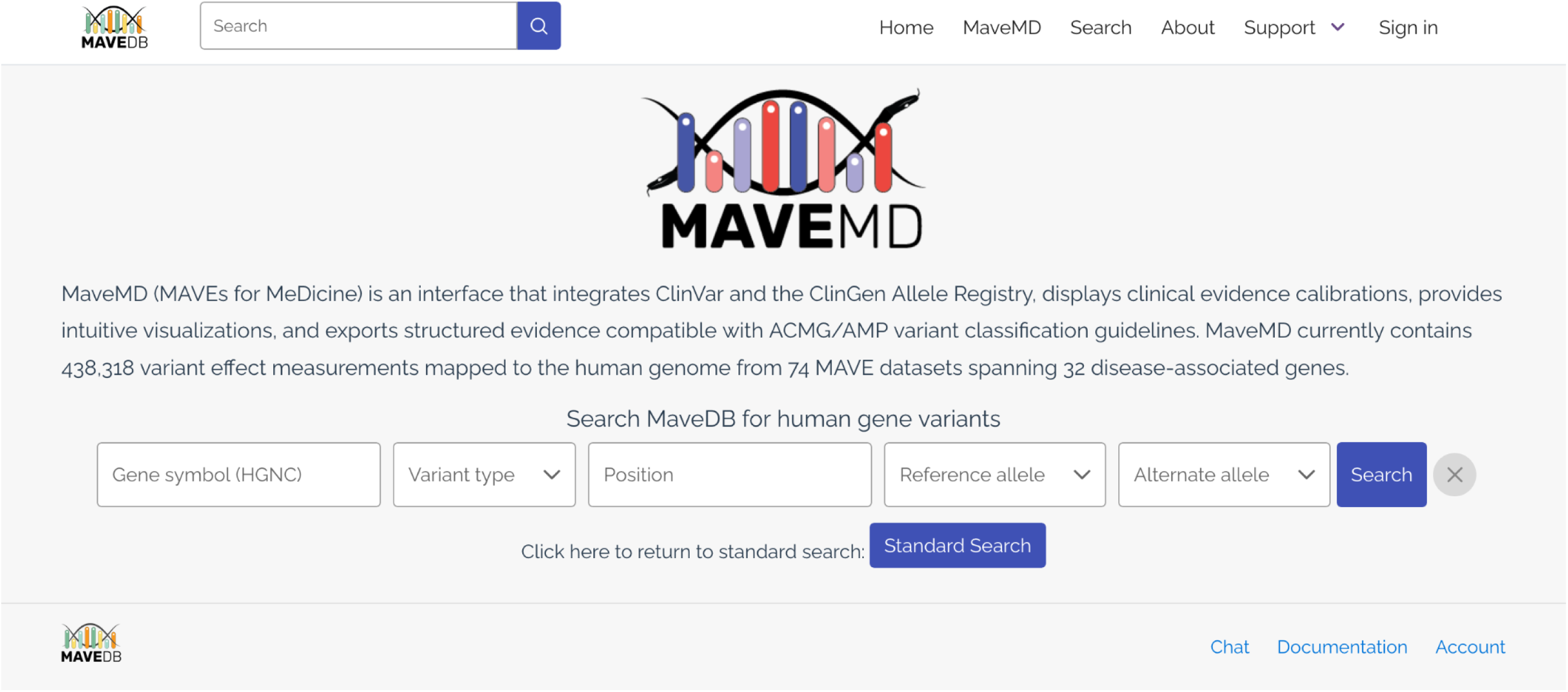
Variant fuzzy search form. This screenshot shows the search interface for the variant fuzzy search implemented in MaveMD. Variant type, reference allele, and alternate allele are dropdown selections, and gene symbol and position are free text. Input to this form is validated and passed to the ClinGen Allele Registry via API to resolve variant search queries.

**Supplemental Figure S3:**
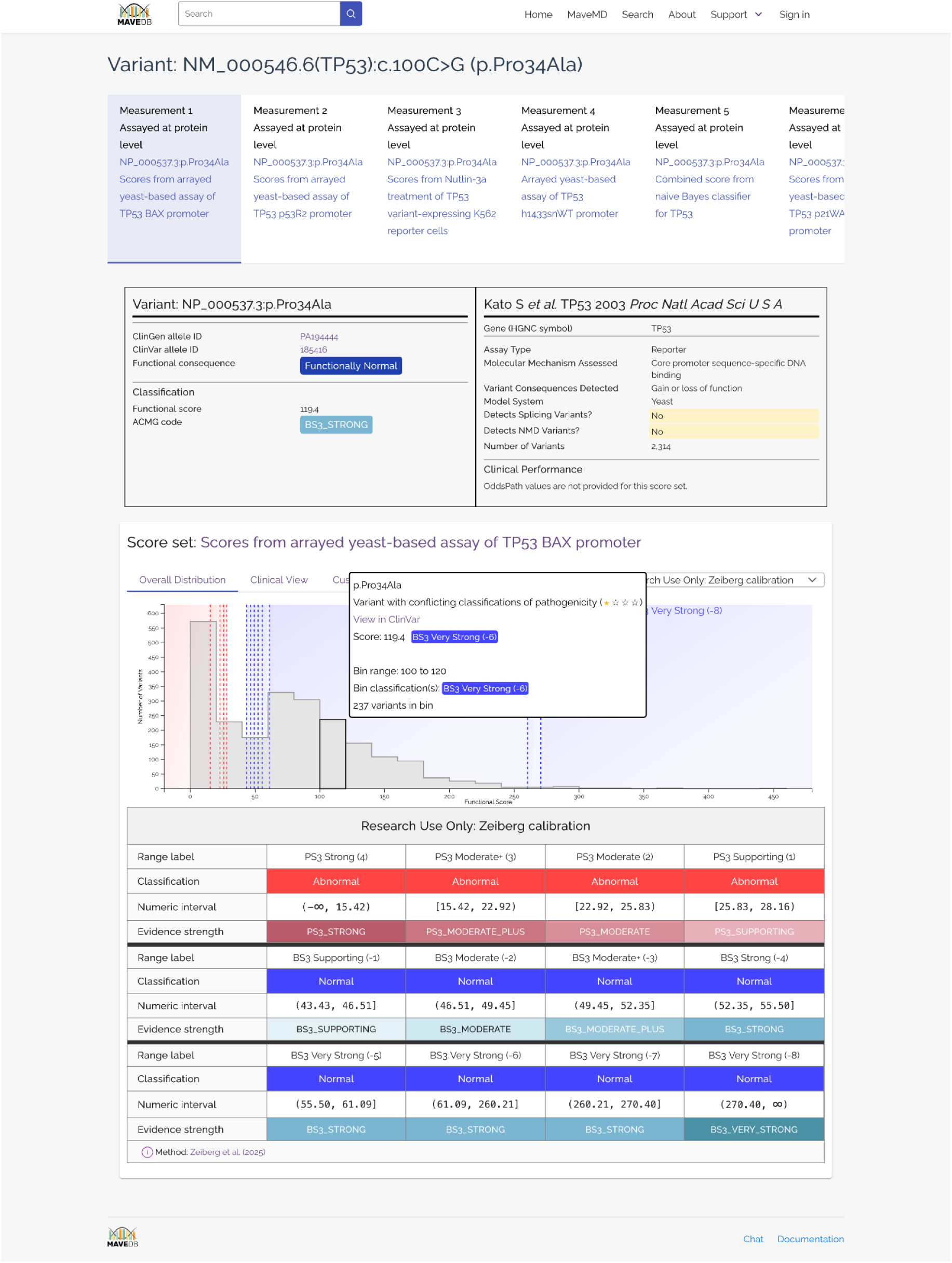
Variant search results page. This screenshot shows the results page for the variant search implemented in MaveMD. Available measurements (assay results) are available at the top of the page and the user can select which one should be used for visualization. The variant is shown in HGVS format, along with alternate identifiers. The variant’s position in the assay score histogram is also highlighted.

**Supplemental Figure S4:**
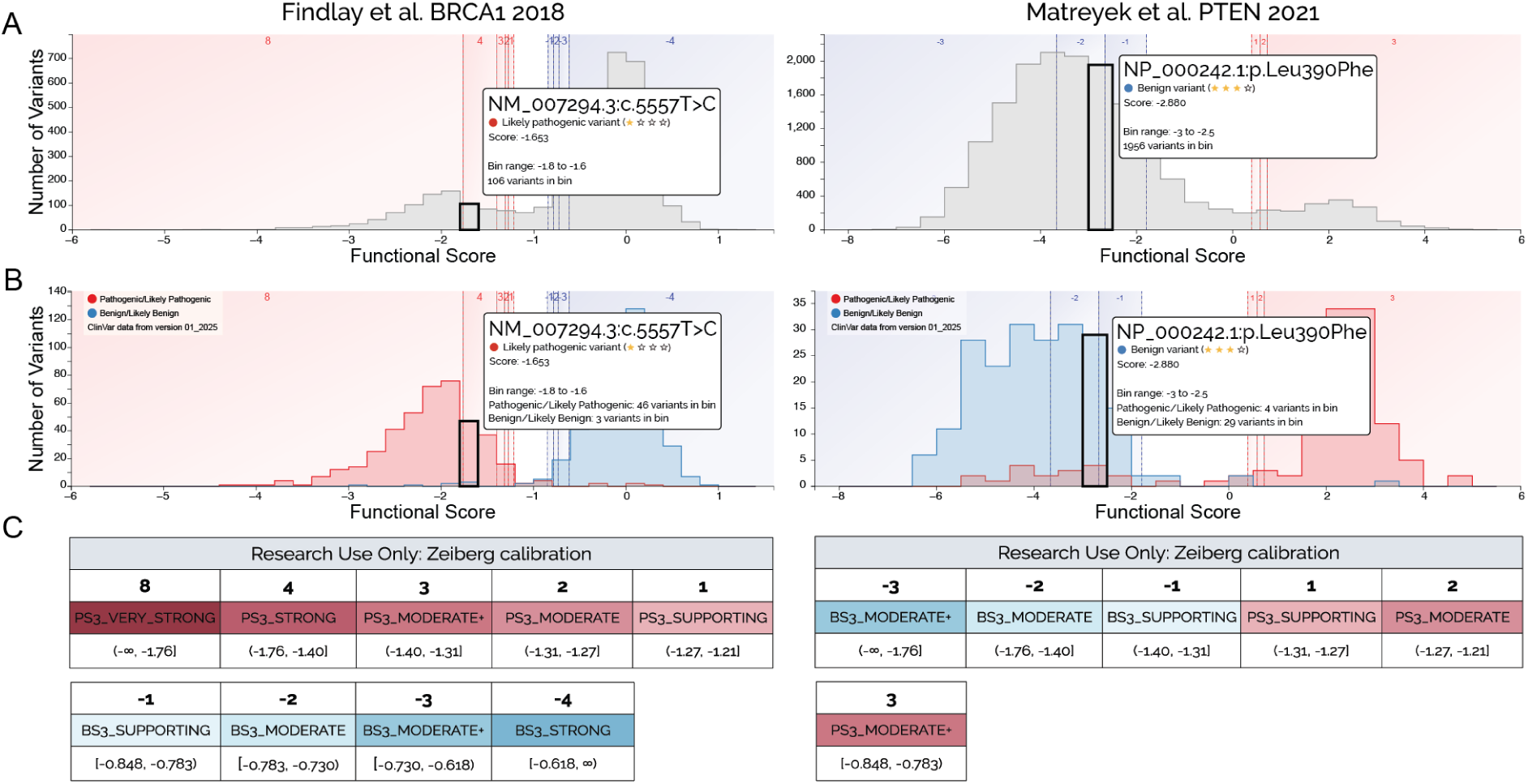
MaveMD support for alternative calibration methods. A) Interactive histograms show the full distribution of variant scores in an assay and the relative position of a selected variant. Vertical bars also denote evidence thresholds. B) Another interactive histogram displays the distribution of variant scores for variants from ClinVar. C) Clinical calibration details and the associated evidence codes and score ranges are shown in a table below the histogram. The histogram shows the calibrations based on the ExCALIBR method^26^.

